# Validation of Algorithms to Identify Small or Large for Gestational Age in the Korean Nationwide Healthcare Database

**DOI:** 10.64898/2026.09.09.26362678

**Authors:** Yongtai Cho, HyunJoo Lim, Bohyun Suh, Yubin Lee, Ju-Young Shin

## Abstract

**Objective:** Small for gestational age (SGA) and large for gestational age (LGA) are critical endpoints in perinatal pharmacoepidemiology, yet gestational age is frequently missing in administrative databases. This study validated algorithms for identifying SGA and LGA in the Korean National Health Information Database (NHID).

**Methods:** We linked the NHID with national vaccination and infant health screening registries (2018–2021) to create a validation cohort of 94,159 pregnancies with reference standard gestational age and birth weight; infants with birth weights below the 10^th^ or above the 90^th^ percentile were considered SGA or LGA, respectively. We evaluated four algorithms: ICD-10 diagnosis codes from infant claims (Method A), maternal claims (Method B), either infant or maternal claims (Method C), and birth weight combined with estimated gestational age (Method D).

**Results:** ICD-10–based algorithms consistently underestimated prevalence, showing high specificity but low sensitivity (<17%). In contrast, Method D demonstrated superior performance: for SGA, sensitivity was 89.3%, specificity 96.8%, and positive predictive value (PPV) 71.6%; for LGA, sensitivity was 77.5%, specificity 98.3%, and PPV 84.7%.

**Conclusion:** SGA and LGA can be identified with reasonable accuracy in the NHID using birth weight and estimated gestational age, whereas diagnosis codes alone can underestimate prevalence.

## INTRODUCTION

Abnormal fetal growth, commonly manifested as small for gestational age (SGA) or large for gestational age (LGA), reflects the intrauterine environment and is strongly associated with adverse neonatal outcomes as well as long-term metabolic and cardiovascular risks in later life.^1,2^ In pharmacoepidemiologic research, SGA and LGA are therefore important indicators of potential teratogenic or fetotoxic effects of medication exposure during pregnancy.^1^

Because pregnant individuals are often excluded from clinical trials, administrative healthcare databases have become an essential resource for evaluating medication safety in pregnancy.^3^ These databases offer large sample sizes and extended follow-up, enabling the assessment of relatively rare exposures and outcomes. However, key clinical parameters, such as birth weight and gestational age at delivery, are frequently unavailable in these databases.^4^ For example, in the Korean National Health Information Database (NHID), birth weight is often captured, whereas gestational age is commonly absent, limiting the direct assessment of SGA and LGA.^5,6^ Therefore, pharmacoepidemiologic studies typically rely on diagnosis codes as proxies for identifying SGA and LGA. The validity of these codes, however, may vary substantially depending on coding practices and reimbursement policies.^4^

Although we recently developed and validated algorithms for estimating gestational age in the Korean NHID,^6,7^ the performance of claims-based algorithms for identifying clinically relevant fetal growth outcomes, such as SGA and LGA, has not been evaluated. The objective of this study was to validate claims-based definitions of SGA and LGA against a reference standard derived from a unique linkage of nationwide databases.

## METHODS

The National Health Information Database (NHID) is a nationwide claims database that captures all reimbursed medical services and demographic information at the patient level for Korea’s compulsory national health insurance program. The NHID was linked to the vaccination registry of the Korea Disease Control and Prevention Agency (KDCA) from January 1, 2018, to December 31, 2021. This registry includes information on gestational age at the time of influenza vaccination, as reported by vaccine recipients.^6^ In addition, the NHID was linked to data from the National Health Screening Program for Infants and Children (NHSPIC), which provides parent-reported information on infant birth weight.^8^ The validation cohort comprised pregnant women who 1) delivered from September 1, 2019 to July 31, 2021 and had linkage to their liveborn infants, 2) had recorded birth weight information, and 3) received influenza vaccination during pregnancy.

We defined the gestational age at delivery by adding the interval between the vaccination date and the delivery date to the reported gestational age at the time of influenza vaccination. The total duration of gestation was then divided by seven and rounded down to obtain gestational age in completed weeks at delivery. Using the calculated gestational age and birth weight, infants were classified as SGA or LGA if their birth weight was below the 10^th^ percentile or above the 90^th^ percentile, respectively, according to infant sex and plurality, based on published population-based reference distributions.^9^

We used four approaches to identify SGA and LGA infants. In Method A, SGA and LGA were identified using diagnosis codes recorded in the infant’s claims within the first 180 days of life. SGA was defined using International Classification of Diseases, 10^th^ Revision (ICD-10) code P05, excluding P05.2, and LGA using ICD-10 code P08, excluding P08.2. In Method B, we identified maternal diagnosis codes recorded from 30 days before delivery through 180 days after delivery. For this method, ICD-10 codes O36.5 and O36.6 were used to identify SGA and LGA, respectively. In Method C, we expanded the definition to include both maternal and infant diagnoses. In Method D, rather than relying on ICD-10 diagnosis codes, SGA and LGA were identified using birth weight in combination with estimated gestational age at delivery, calculated using a previously validated method.^6,7^ The present study is distinct because it evaluates the validity of algorithms for identifying SGA and LGA, including both diagnosis-code-based definitions and gestational-age-based definitions, against a reference standard.

We compared demographic, healthcare utilization, and clinical characteristics between individuals included in the validation dataset and all pregnancies linked to liveborn infants during the study period (**Supplementary Table 1**). Maternal comorbidities were assessed from 90 days before the last menstrual period (LMP) through delivery, with the exception of preeclampsia/eclampsia, which was assessed from 140 days after the LMP through delivery. Differences between the two groups were described using standardized mean differences (SMD).

**Table 1.** Performance metrics for claims-based methods for identifying small and large for gestational age.

|  | **Sensitivity,**<br>**% (95% CI)** | **Specificity,**<br>**% (95% CI)** | **PPV,**<br>**% (95% CI)** | **NPV,**<br>**% (95% CI)**<sup>256</sup> |
| --- | --- | --- | --- | --- |
| **Main analysis** |  |  |  |  |
| **Small for gestational age**<sup>258</sup> |  |  |  |  |
| Method A: Code on infant claims | 5.6 (5.1–6.1) | 99.9 (99.8–99.9) | 76.8 (73.4–80.3) | 92.1 (92.0–92.3) |
| Method B: Code on maternal claims | 13.0 (12.2–13.7) | 98.9 (98.8–99.0) | 52.2 (50.0–42.4) | 92.6 (92.5–92.8) |
| Method C: Code on infant or maternal claims | 16.4 (15.6–17.3) | 98.8 (98.7–98.8) | 54.6 (52.6–56.6) | 92.9 (92.7–93.1) |
| Method D: Based on estimated gestational age | 89.3 (88.6–90.0) | 96.8 (96.7–96.9) | 71.6 (70.8–72.5) | 99.0 (98.9–99.1) |
| **Large for gestational age** |  |  |  |  |
| Method A: Code on infant claims | 8.4 (7.8–8.9) | 99.9 (99.9–99.9) | 92.1 (90.4–93.8) | 90.0 (89.8–90.2) |
| Method B: Code on maternal claims | 3.6 (3.2–3.9) | 99.8 (99.7–99.8) | 64.4 (60.3–68.4) | 89.6 (89.4–89.8) |
| Method C: Code on infant or maternal claims | 10.8 (10.2–11.4) | 99.7 (99.6–99.7) | 80.4 (78.2–82.5) | 90.3 (90.1–90.5) |
| Method D: Based on estimated gestational age | 77.5 (76.7–78.3) | 98.3 (98.2–98.4) | 84.7 (84.0–85.5) | 97.3 (97.2–97.4) |
| **Sensitivity analysis: validation set weighted to represent the general population**<sup>265</sup> |  |  |  |  |
| **Small for gestational age** |  |  |  |  |
| Method A: Code on infant claims | 6.1 (5.5–6.6) | 99.8 (99.8–99.9) | 75.4 (71.9–78.8) | 92.2 (92.0–92.3) |
| Method B: Code on maternal claims | 13.4 (12.6–14.2) | 98.9 (98.8–99.0) | 52.2 (50.0–54.4) | 92.7 (92.5–92.8) |
| Method C: Code on infant or maternal claims | 17.2 (16.3–18.0) | 98.7 (98.7–98.8) | 55.1 (53.1–57.1) | 92.9 (92.8–93.1) |
| Method D: Based on estimated gestational age | 88.4 (87.7–89.1) | 96.8 (96.7–97.0) | 71.7 (70.7–72.6) | 98.9 (98.9–99.0) |
| **Large for gestational age**<sup>268</sup> |  |  |  |  |
| Method A: Code on infant claims | 8.6 (8.0–9.1) | 99.9 (99.9–99.9) | 92.9 (91.2–94.6) | 90.0 (89.8–90.2) |
| Method B: Code on maternal claims | 3.6 (3.3–4.0) | 99.8 (99.7–99.8) | 65.4 (61.4–69.4) | 89.5 (89.3–89.7) |
| Method C: Code on infant or maternal claims | 11.1 (10.5–11.7) | 99.7 (99.7–99.7) | 81.4 (79.3–83.5) | 90.3 (90.1–90.4) |
| Method D: Based on estimated gestational age | 78.2 (77.4–79.0) | 98.0 (98.0–98.1) | 82.9 (82.1–83.7) | 97.4 (97.3–97.5) |

Abbreviations: CI, confidence interval; NPV, negative predictive value; PPV, positive predictive value

The performance of each identification method was evaluated using four metrics: sensitivity, specificity, positive predictive value (PPV), and negative predictive value (NPV). 95% confidence intervals (CIs) were estimated assuming a binomial distribution. As a sensitivity analysis, we applied inverse probability of selection weights derived from all measured characteristics to improve representativeness of the general population.^10^ To examine whether algorithm performance varied by maternal age, income level, infant sex, gestational age, and prenatal care utilization index,^11^ we calculated performance metrics stratified by each subgroup using the best-performing algorithm. All statistical analyses were performed using SAS 9.4 (SAS Institute Inc.). This study used data provided by the National Health Insurance Service (NHIS) under data access approval No. NHIS-2022-1-749. The study was granted exemption from Institutional Review Board (IRB) review by the Sungkyunkwan University IRB, South Korea (2025-12-054).

## RESULTS

Among 403,031 pregnancies resulting in live birth during the inclusion period, 391,343 (97.1%) had recorded birth weight. Of those, 94,159 (24.1%) received influenza vaccination during pregnancy and thus had gestational age information available, and were therefore included in the validation cohort. Compared with the overall population of live births, the validation cohort had fewer deliveries occurring in summer and more occurring in winter. The validation cohort also had a higher proportion of pregnancies receiving adequate-plus prenatal care (**Figure 1**). All characteristics were well balanced after applying inverse probability weighting in the sensitivity analysis (**Supplementary Table 2**).

**Figure 1.**
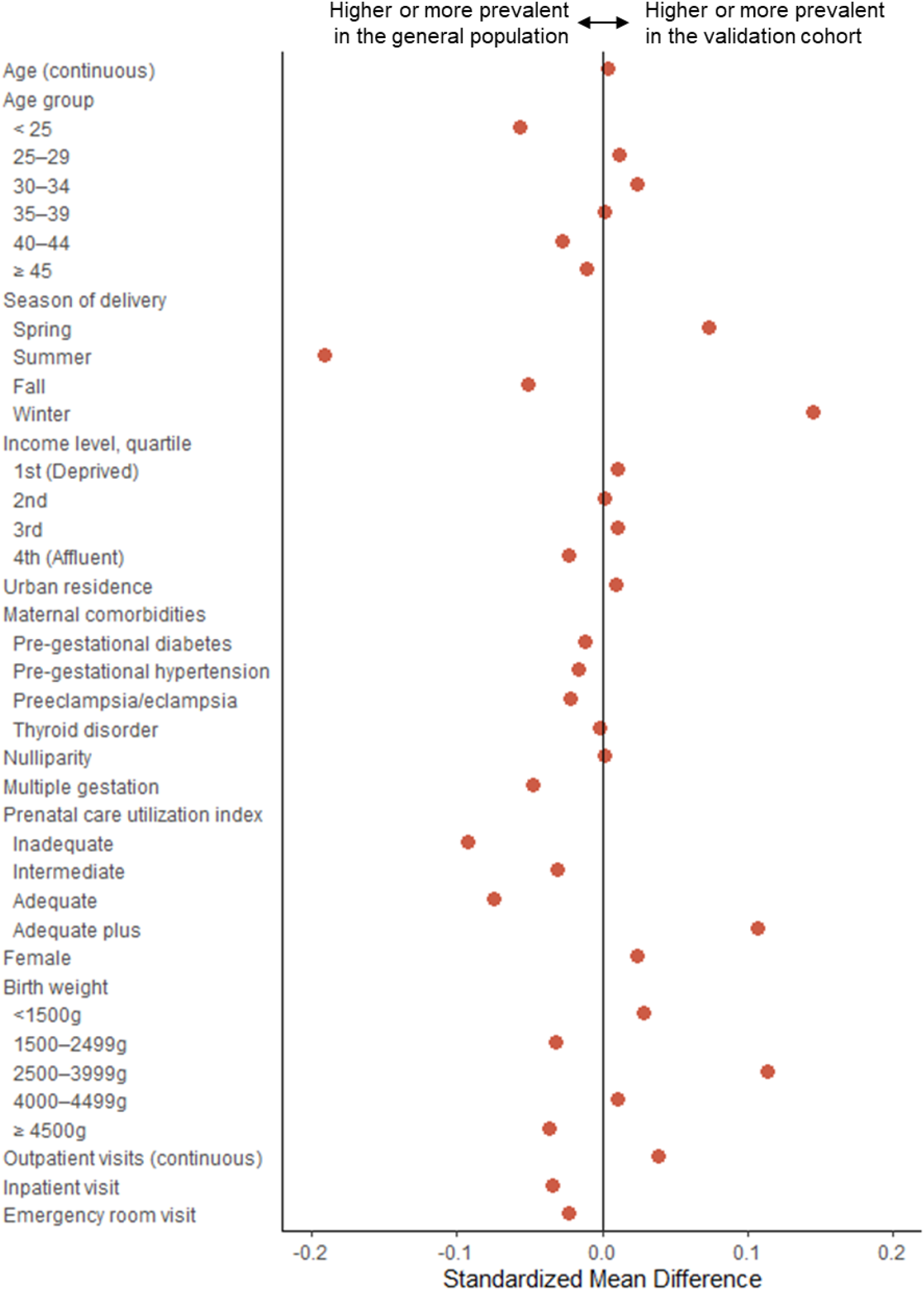
Comparison of the validation cohort versus all live births during the study period *All characteristics were measured as binary variables unless indicated otherwise.

For both SGA and LGA, ICD-10–based approaches (Methods A–C) markedly underestimated prevalence, with very high specificity but low sensitivity. Adding maternal diagnosis codes modestly improved sensitivity for SGA (5.6% to 16.4%) and LGA (8.4% to 10.8%), at the expense of reduced PPV (SGA: 76.8% to 54.6%, LGA: 92.1% to 80.4%). In contrast, identifying SGA and LGA using estimated gestational age (Method D) demonstrated the best overall performance. For SGA, this approach achieved a sensitivity of 89.3%, specificity of 96.8%, PPV of 71.6%, and NPV of 99.0%. For LGA, corresponding performance metrics were 77.5% sensitivity, 98.3% specificity, 84.7% PPV, and 97.3% NPV. Results were consistent after applying inverse probability of selection weights (**Table 1**).

When stratified by subgroups, Method D showed lower performance among infants born at earlier gestational ages and among pregnancies with inadequate prenatal care. For SGA, PPV was lower for preterm than term births (< 29 weeks: 36.0% vs. ≥ 37 weeks: 72.9%), while for LGA, sensitivity was lower in preterm births (< 29 weeks: 16.6% vs. ≥ 37 weeks: 82.2%). Inadequate prenatal care was associated with lower sensitivity for SGA (47.7% vs. 91.3%) and lower PPV for LGA (19.3% vs. 81.9%) compared with adequate-plus care (**Supplementary Tables 3** and **4**).

## DISCUSSION

This validation study demonstrated that SGA and LGA can be identified with reasonable reliability in the NHID when birth weight information is combined with estimated gestational age, whereas definitions based solely on ICD-10 diagnosis codes tend to underestimate the prevalence of both conditions. Although claims-based definitions of SGA and LGA have been validated in the United States and European countries,^12,13^ establishing their transportability across healthcare systems is essential given differences in population characteristics and clinical practice patterns.^4^ To our knowledge, the validity of these diagnoses has not previously been assessed in the NHID.

We observed heterogeneity in the performance of Method D across patient subgroups. For SGA, the PPV was lower among preterm births, consistent with findings from algorithms developed in the Medicaid database.^13^ In contrast, for LGA, PPV remained relatively stable while sensitivity decreased among preterm births. An opposite pattern was observed among pregnancies with inadequate prenatal care, with reduced sensitivity for SGA and lower PPV for LGA compared with those receiving adequate-plus care. This may be explained by greater uncertainty in gestational age estimation, as Method D relies on prenatal care information (e.g., timing of ultrasound examinations).^6^ If gestational age is underestimated in this population, infants may appear relatively larger for their assigned gestational age, increasing false negative classifications for SGA and false positive classifications for LGA. Potential differential misclassification of SGA and LGA should be considered when making comparisons across populations with differing characteristics.

Our results should be interpreted in light of several limitations. The validation cohort in this study was restricted to individuals who received influenza vaccination during pregnancy, who may differ from the general pregnant population in healthcare utilization patterns. To address this, we conducted a sensitivity analysis using inverse probability of selection weighting based on demographic, clinical, and healthcare utilization characteristics, which yielded consistent results. Furthermore, because the study was restricted to live births, the performance of these algorithms could not be evaluated for pregnancies ending earlier. Finally, given differences in coding practices and reimbursement policies, the generalizability of these methods to settings outside Korea may be limited.

In conclusion, when birth weight data are available, SGA and LGA can be identified with reasonable accuracy in the NHID. Population characteristics should be considered when using SGA or LGA as study outcomes, as algorithm performance varied across population subgroups. Future studies evaluating medication effects on fetal growth outcomes should account for the potential impact of outcome misclassification in the study design and interpretation.

## Data Availability

Data generated and/or analyzed during the current study cannot be shared publicly due to the data-sharing policy of the National Health Insurance Service (NHIS) of Korea, governed by Article 18 of the Personal Information Protection Act (Limitation to Out-of-Purpose Use and Provision of Personal Information available at https://elaw.klri.re.kr/kor_service/lawView.do?hseq=53044&lang=ENG). However, the data are available from the NHIS (study identifier: NHIS-2022-1-740) on reasonable request for researchers who meet the criteria for access to confidential data (https://www.data.go.kr/en/tcs/eds/selectCoreDataView.do?coreDataInsttCode=B551182&coreDataSn=1&searchCondition2=coreDataNmEn&searchKeyword2=).

## Notes

**Funding:** This work was supported by a grant [RS-2026-25475077] from the National Research Foundation of Korea, which is funded by the Korean government (Ministry of Science and Information and Communication Technology).

**Conflict of interest:** Ju-Young Shin received grants from the Ministry of Food and Drug Safety, the Ministry of Health and Welfare, the National Research Foundation of Korea and pharmaceutical companies, including Pfizer, UCB, and Yuhan, outside the submitted work.

### Competing Interest Statement

Ju-Young Shin received grants from the Ministry of Food and Drug Safety, the Ministry of Health and Welfare, the National Research Foundation of Korea and pharmaceutical companies, including Pfizer, UCB, and Yuhan, outside the submitted work.

### Author Declarations

The study was granted exemption from Institutional Review Board (IRB) review by the Sungkyunkwan University IRB, South Korea (2025-12-054). Under Article 13(1)3 of the Enforcement Rule of the Korean Bioethics and Safety Act, research using existing data or documents that does not collect or record personally identifiable information is eligible for exemption from IRB review. Because this study involved secondary analysis of existing de-identified data without any contact with participants, individual informed consent was not obtained.

