## Supplementary Table for "Validation of Algorithms to Identify Small or Large for Gestational Age in the Korean Nationwide Healthcare Database"


| Table of Contents | Page |
|---|---|
| **Supplementary Table 1.** ICD-10 codes used to define study variables | 2 |
| **Supplementary Table 2.** Maternal and neonatal characteristics of the validation cohort and all pregnancies resulting in live birth during the study period | 3 |
| **Supplementary Table 3.** Performance metrics for the best performing claims-based method for identifying small for gestational age, stratified by subgroups | 4 |
| **Supplementary Table 4.** Performance metrics for the best performing claims-based method for identifying large for gestational age, stratified by subgroups | 5 |

**Supplementary Table 1.** ICD-10 codes used to define study variables

| Categories | Codes |
|---|---|
| **Outcome** | **ICD-10 codes** |
| Small for gestational age | P05 (excl. P05.2), O36.5 (mother only) |
| Large for gestational age | P08 (excl. P08.2), O36.6 (mother only) |
| **Covariate** | **ICD-10 codes** |
| Pre-gestational diabetes | E10–E14, O24.0–O24.3 |
| Pre-gestational hypertension | O10, I10–I15 |
| Preeclampsia/eclampsia | O11, O14, O15 |
| Thyroid disorder | E02, E03, E05, E06 |

Abbreviation: ICD-10, International Classification of Diseases, 10th Revision

**Supplementary Table 2.** Maternal and neonatal characteristics of the validation cohort and all pregnancies resulting in live birth during the study period

| | Before weighting, n (%) | | | After weighting, n(%) | | |
|---|---|---|---|---|---|---|
| | **Validation cohort** | **All pregnancies** | **SMD** | **Validation cohort** | **All pregnancies** | **SMD** |
| Total | 94,159 (100) | 403,031 (100) | NA | 94,048 (100) | 403,031 (100) | NA |
| Age, mean (SD) | 33.1 (4.3) | 33.1 (4.5) | 0.004 | 33.1 (4.4) | 33.1 (4.5) | -0.001 |
| Age group | | | | | | |
| < 25 | 2,352 (2.5) | 13,987 (3.5) | -0.057 | 3,172 (3.4) | 13,987 (3.5) | -0.005 |
| 25–29 | 16,236 (17.2) | 67,712 (16.8) | 0.012 | 15,839 (16.8) | 67,712 (16.8) | 0.001 |
| 30–34 | 39,867 (42.3) | 165,954 (41.2) | 0.024 | 38,946 (41.4) | 165,954 (41.2) | 0.005 |
| 35–39 | 29,721 (31.6) | 126,929 (31.5) | 0.002 | 29,505 (31.4) | 126,929 (31.5) | -0.003 |
| 40–44 | 5,821 (6.2) | 27,561 (6.8) | -0.027 | 6,386 (6.8) | 27,561 (6.8) | -0.002 |
| ≥ 45 | 162 (0.2) | 888 (0.2) | -0.011 | 201 (0.2) | 888 (0.2) | -0.001 |
| Season of delivery[a] | | | | | | |
| Spring | 27,654 (29.4) | 105,210 (26.1) | 0.073 | 24,592 (26.1) | 105,210 (26.1) | 0.001 |
| Summer | 12,973 (13.8) | 84,502 (21.0) | -0.191 | 19,551 (20.8) | 84,502 (21.0) | -0.004 |
| Fall | 22,857 (24.3) | 108,488 (26.9) | -0.051 | 25,311 (26.9) | 108,488 (26.9) | 0.000 |
| Winter | 30,675 (32.6) | 104,831 (26.0) | 0.145 | 19,377 (20.6) | 104,831 (26.0) | 0.003 |
| Income level, quartile | | | | | | |
| 1st (Deprived) | 19,746 (21.0) | 82,938 (20.6) | 0.010 | 19,377 (20.6) | 82,938 (20.6) | 0.001 |
| 2nd | 21,905 (23.3) | 93,663 (23.2) | 0.001 | 21,890 (23.3) | 93,663 (23.2) | 0.001 |
| 3rd | 32,278 (34.3) | 136,004 (33.7) | 0.011 | 31,706 (33.7) | 136,004 (33.7) | -0.001 |
| 4th (Affluent) | 20.230 (21.5) | 90,426 (22.4) | -0.023 | 21,075 (22.4) | 90,426 (22.4) | -0.001 |
| Urban residence | 66,378 (70.5) | 282,403 (70.1) | 0.009 | 66,106 (70.3) | 282,403 (70.1) | 0.005 |
| Maternal comorbidities | | | | | | |
| Pregestational diabetes | 2,266 (2.4) | 10,483 (2.6) | -0.012 | 2,463 (2.6) | 10,483 (2.6) | 0.001 |
| Pregestational hypertension | 1,292 (1.4) | 6,288 (1.6) | -0.016 | 1,482 (1.6) | 6,288 (1.6) | 0.001 |
| Preeclampsia/eclampsia | 611 (0.6) | 3,389 (0.8) | -0.022 | 807 (0.9) | 3,389 (0.8) | 0.002 |
| Thyroid disorder | 13,992 (14.9) | 60.224 (14.9) | -0.002 | 14,164 (15.1) | 60.224 (14.9) | 0.003 |
| Nulliparity | 50,978 (54.1) | 217,893 (54.1) | 0.002 | 51,464 (54.7) | 217,893 (54.1) | 0.013 |
| Multiple gestation | 3,537 (3.8) | 19,078 (4.7) | -0.048 | 4,365 (4.6) | 19,078 (4.7) | -0.004 |
| Prenatal care utilization index | | | | | | |
| Inadequate | 994 (1.1) | 8,960 (2.2) | -0.092 | 2,006 (2.1) | 8,960 (2.2) | -0.006 |
| Intermediate | 1,436 (1.5) | 7,786 (1.9) | -0.031 | 1,795 (1.9) | 7,786 (1.9) | -0.002 |
| Adequate | 16,589 (17.6) | 82,651 (20.5) | -0.074 | 19,117 (20.3) | 82,651 (20.5) | -0.004 |
| Adequate plus | 75,139 (79.8) | 303,634 (75.3) | 0.107 | 71,131 (75.6) | 303,634 (75.3) | 0.007 |
| Female | 45,712 (48.5) | 190,746 (47.3) | 0.024 | 44,673 (47.5) | 190,746 (47.3) | 0.003 |
| Birth weight | | | | | | |
| <1500g | 742 (0.8) | 2,256 (0.6) | 0.028 | 465 (0.5) | 2,256 (0.6) | -0.009 |
| 1500–2499g | 4,886 (5.2) | 23,856 (5.9) | -0.032 | 5,428 (5.8) | 23,856 (5.9) | -0.006 |
| 2500–3999g | 85,845 (91.2) | 353,521 (87.7) | 0.113 | 85,403 (90.8) | 365,209 (90.6) | 0.007 |
| 4000–4499g | 2,434 (2.6) | 9,788 (2.4) | 0.010 | 2,296 (2.4) | 9,788 (2.4) | 0.001 |
| ≥ 4500g | 243 (0.3) | 1,922 (0.5) | -0.036 | 457 (0.5) | 1,922 (0.5) | 0.001 |
| Missing[b] | 0 (0.0) | 11,688 (2.9) | NA | 0 (0.0) | 0 (0.0) | NA |
| No. outpatient visits during pregnancy, mean (SD) | 19.7 (7.0) | 19.4 (7.4) | 0.039 | 19.5 (7.0) | 19.4 (7.4) | 0.009 |
| Inpatient visit during pregnancy | 14,064 (14.9) | 65,225 (16.2) | -0.034 | 15,105 (16.1) | 65,225 (16.2) | -0.003 |
| Emergency room visit during pregnancy | 8,235 (8.7) | 37,904 (9.4) | -0.023 | 8,782 (9.3) | 37,904 (9.4) | -0.002 |

Abbreviations: SD, standard deviation; SMD, standardized mean difference

[a] Defined as follows: Spring, March to May; summer, July to August; fall, September to November; Winter, December to February

[b] Missing values were imputed median birth weight for the inverse probability weight model

| Small for gestational age | Sensitivity, % (95% CI) | Specificity, % (95% CI) | PPV, % (95% CI) | NPV, % (95% CI) |
|---|---|---|---|---|
| **Maternal age, years** | | | | |
| < 25 | 80.2 (75.0–85.4) | 96.3 (95.5–97.1) | 70.0 (64.4–75.6) | 97.9 (97.2–98.5) |
| 25–29 | 89.8 (88.2–91.4) | 96.6 (96.3–96.9) | 71.0 (68.8–73.1) | 99.0 (98.9–99.2) |
| 30–34 | 90.0 (89.0–91.0) | 97.1 (96.9–97.2) | 73.2 (71.8–74.6) | 99.1 (99.0–99.2) |
| 35–39 | 89.1 (87.8–90.3) | 96.7 (96.4–96.9) | 70.1 (68.5–71.7) | 99.0 (98.9–99.1) |
| 40–44 | 88.3 (85.5–91.1) | 96.7 (96.2–97.1) | 71.5 (68.0–75.1) | 98.9 (98.6–99.2) |
| ≥ 45 | 88.9 (74.4–100) | 96.5 (93.5–99.5) | 76.2 (58.0–94.4) | 98.6 (96.6–100) |
| **Income level, quartile** | | | | |
| 1st (Deprived) | 88.4 (86.8–89.9) | 96.6 (96.4–96.9) | 71.1 (69.2–73.1) | 98.9 (98.7–99.0) |
| 2nd | 88.9 (87.5–90.4) | 96.7 (96.4–96.9) | 71.5 (69.7–73.3) | 98.9 (98.8–99.1) |
| 3rd | 89.3 (88.2–90.5) | 96.9 (96.7–97.1) | 71.7 (70.2–73.3) | 99.0 (98.9–99.1) |
| 4th (Affluent) | 90.6 (89.1–92.0) | 97.0 (96.7–97.2) | 72.2 (70.3–74.2) | 99.2 (99.0–99.3) |
| **Infant sex** | | | | |
| Female | 89.6 (88.6–90.6) | 96.7 (96.6–96.9) | 71.2 (69.9–72.5) | 99.0 (98.9–99.1) |
| Male | 89.0 (88.1–90.0) | 96.9 (96.7–97.0) | 72.1 (70.8–73.3) | 99.0 (98.9–99.1) |
| **Gestational age, weeks** | | | | |
| < 29 | 100 (100–100) | 93.1 (89.9–96.4) | 36.0 (17.2–54.8) | 100 (100–100) |
| 29–31 | 94.4 (83.9–100) | 89.6 (86.3–93.0) | 34.7 (21.4–48.0) | 99.6 (98.9–100) |

**Supplementary Table 3.** Performance metrics for the best performing claims-based method[a] for identifying small for gestational age, stratified by subgroups

| | | | | |
|---|---|---|---|---|
| 32–34 | 89.8 (85.2–94.4) | 91.8 (90.3–93.4) | 60.7 (54.6–66.8) | 98.5 (97.7–99.2) |
| 35–36 | 93.5 (91.2–95.7) | 94.9 (94.2–95.5) | 63.6 (60.0–67.2) | 99.3 (99.1–99.6) |
| ≥ 37 | 89.0 (88.2–89.7) | 97.0 (96.9–97.2) | 72.9 (72.0–73.8) | 99.0 (98.9–99.1) |
| **Prenatal care utilization index** | | | | |
| Inadequate | 47.7 (38.2–57.1) | 98.2 (97.3–99.1) | 76.1 (65.9–86.3) | 94.0 (92.4–95.5) |
| Intermediate | 68.0 (58.8–77.3) | 97.9 (97.1–98.7) | 70.2 (61.0–79.5) | 97.7 (96.9–98.5) |
| Adequate | 84.6 (82.7–86.6) | 97.6 (97.4–97.9) | 75.4 (73.2–77.6) | 98.7 (98.5–98.8) |
| Adequate plus | 91.3 (90.6–92.0) | 96.6 (96.5–96.7) | 70.9 (69.9–71.9) | 99.2 (99.1–99.3) |

Abbreviations: CI, confidence interval; NPV, negative predictive value; PPV, positive predictive value

[a] Identifying small for gestational age by using estimated gestational age and birth weight information

| Large for gestational age | Sensitivity, % (95% CI) | Specificity, % (95% CI) | PPV, % (95% CI) | NPV, % (95% CI) |
|---|---|---|---|---|
| **Maternal age, years** | | | | |
| < 25 | 74.1 (67.9–80.3) | 95.7 (94.9–96.6) | 60.9 (54.6–67.1) | 97.6 (97.0–98.3) |
| 25–29 | 76.2 (74.1–78.3) | 98.5 (98.3–98.7) | 85.4 (83.6–87.2) | 97.3 (97.1–97.6) |
| 30–34 | 77.9 (76.6–79.1) | 98.6 (98.5–98.7) | 86.4 (85.2–87.5) | 97.5 (97.3–97.6) |
| 35–39 | 78.0 (76.6–79.4) | 98.2 (98.1–98.4) | 84.9 (83.6–86.2) | 97.2 (97.0–97.4) |
| 40–44 | 76.5 (73.6–79.5) | 97.5 (97.1–97.9) | 82.6 (79.9–85.4) | 96.4 (95.9–96.9) |
| ≥ 45 | 84.6 (65.0–100) | 96.6 (93.8–99.5) | 68.8 (46.0–91.5) | 98.6 (96.7–100) |
| **Income level, quartile** | | | | |
| 1st (Deprived) | 76.2 (74.4–77.9) | 97.8 (97.6–98.0) | 81.2 (79.5–82.9) | 97.0 (96.8–97.3) |
| 2nd | 77.9 (76.3–79.6) | 98.3 (98.1–98.5) | 84.9 (83.4–86.4) | 97.3 (97.1–97.5) |
| 3rd | 78.6 (77.2–79.9) | 98.6 (98.4–98.7) | 86.7 (85.5–87.9) | 97.5 (97.3–97.7) |
| 4th (Affluent) | 76.6 (74.8–78.4) | 98.4 (98.3–98.6) | 85.2 (83.6–86.8) | 97.3 (97.0–97.5) |
| **Infant sex** | | | | |
| Female | 78.3 (77.2–79.5) | 98.4 (98.3–98.5) | 85.0 (83.9–86.0) | 97.5 (97.3–97.6) |
| Male | 76.7 (75.6–77.8) | 98.3 (98.1–98.4) | 84.5 (83.5–85.6) | 97.2 (97.0–97.3) |
| **Gestational age, weeks** | | | | |
| < 29 | 16.6 (11.0–22.2) | 100 (100–100) | 100 (100–100) | 34.1 (27.8–40.5) |
| 29–31 | 20.0 (13.0–27.0) | 99.5 (98.5–100) | 96.2 (88.8–100) | 66.8 (61.5–72.1) |

**Supplementary Table 4.** Performance metrics for the best performing claims-based method[a] for identifying large for gestational age, stratified by subgroups

| | | | | |
|---|---|---|---|---|
| 32–34 | 33.5 (27.3–39.7) | 98.6 (97.9–99.3) | 82.4 (74.6–90.2) | 88.2 (86.4–90.0) |
| 35–36 | 54.3 (50.5–58.2) | 97.8 (97.4–98.2) | 76.7 (72.8–80.6) | 94.1 (93.5–94.8) |
| ≥ 37 | 82.2 (81.4–82.9) | 98.3 (98.3–98.4) | 85.1 (84.4–85.9) | 98.0 (97.9–98.1) |
| **Prenatal care utilization index** | | | | |
| Inadequate | 81.8 (73.2–90.4) | 71.3 (68.4–74.3) | 19.3 (15.0–23.6) | 97.9 (96.8–99.0) |
| Intermediate | 88.4 (83.3–93.4) | 94.2 (93.0–95.5) | 64.9 (58.5–71.4) | 98.5 (97.9–99.2) |
| Adequate | 85.5 (83.8–87.2) | 97.9 (97.7–98.1) | 81.5 (79.6–83.3) | 98.4 (98.2–98.6) |
| Adequate plus | 85.8 (84.2–87.4) | 97.9 (97.7–98.1) | 81.9 (80.2–83.6) | 98.4 (98.3–98.6) |

Abbreviations: CI, confidence interval; NPV, negative predictive value; PPV, positive predictive value

[a] Identifying large for gestational age by using estimated gestational age and birth weight information
